# Precise Language Responses Challenge Easy Rating Scales - Comparing Clinicians’ and Respondents’ Views

**DOI:** 10.1101/2022.04.23.22274132

**Authors:** Sverker Sikström, Alfred Pålsson Höök, Oscar Kjell

## Abstract

**Background:** Closed-ended rating scales are the most used response format for researchers and clinicians to quantify mental states, whereas in natural contexts people communicate with natural language. The reason for using such scales is that they are typically argued to be more precise in measuring mental constructs, whereas the respondents’ views as to what best communicates mental states are frequently ignored.

**Methods:** We assessed respondents’ (*N* = 304) degree of depression using rating scales, descriptive words, selected words, and free text responses and probed the respondents and clinicians (*N* = 40) for their attitudes to the response formats across twelve dimensions related to the precision of communicating their mental states and the ease of responding.

**Results:** Respondents found free text to be more precise (e.g., *precision d*’ = .88, *elaboration d*’ = 2.0) than rating scales, whereas rating scales were rated as easier to respond to (e.g., *easier d*’ = – .67, *faster d*’ = –1.13). Respondents preferred the free text responses to a greater degree than rating scales compared to clinicians.

**Conclusions:** These finding supports the idea that future assessment of mental health can be aided by computational method based on text data.

## Introduction

Language is the natural way of communicating mental states among most groups in our society. The notable exceptions are behavioural scientists, where close-ended rating scales are the dominating method to examine respondents’ states of mind. For example, in social psychology articles there are typically 20 scales in each article, where 87% of them can be categorized as rating scales (Flake, Pek, & Hehman, 2017). The most likely reason for this is that rating scales are easy to collect and quantify and have been found to have reasonably high validity with regard to the construct being measured.

The argument that rating scales should be preferred over open-ended responses due to high validity has recently been challenged by interdisciplinary studies based on open-ended questions and progress in natural language processing (NLP). Question-based computational language assessments (QCLA), where respondents are asked one or more open-ended questions and the responses are analysed with NLP and machine learning methods, has become a promising alternative to rating scales. For example, Kjell et al. (2019) showed that descriptive word responses concerning harmony in life, satisfaction with life, worry, and depression can be quantified so that they correlate well with corresponding, well-established rating scales. Furthermore, similar constructs, such as depression and anxiety, tend to correlate strongly with rating scales, whereas they are better differentiated by QCLA (Kjell et al, 2019). In addition, pictures portraying facial expressions are more accurately differentiated when described by participants using descriptive words compared with rating scales (Kjell et al., 2019).

These findings are particularly interesting as opened-ended questions evaluated by humans have not shown to increase the validity of mental health assessment. Friborg and Rosenvinge (2013) let students and assistants evaluate the responses from open-ended questions of mental health, and found that their evaluations did not increase the statistical prediction of mental health one year later, over and beyond closed-ended rating scales. Similar, data collected with telephone interviews on the degree of drinking, showed larger extent of drinking for closed-ended responses compared to human evaluations of opened-ended responses (Ivis et al, 1997). The possibility that NLP methods can make contribution, in addition to human performance, needs to be investigated in relation to these studies.

The opportunities for using QCLA have been increased by recent progress in computational methods. For example, BERT (Bidirectional Encoder Representations from Transformers; Devlin et al., 2019) is a language model based on deep learning neural networks using transformers (Vaswani et al., 2017) and has greatly improved the accuracy of NLP models. Recently, BERT has been used to predict rating scales with an accuracy (*r* = .85) that challenges the theoretical upper limits of test-retest reliability (Kjell et al., 2021). This was done using multiple free texts and descriptive word responses regarding harmony in life and satisfaction with life as inputs. Further, Sikström et al. (in progress) showed that a combination of word responses and rating scales diminishes the number of misses in diagnosis (keeping false alarms constant) of depression and anxiety by approximately half compared to state-of-the-art rating scales (the Patient Health Questionnaire, PHQ-9, [Kroenke, Spitzer, Williams 2001] and the Generalized Anxiety Disorder Scale, GAD-7 [Spitzer, Kroenke, Williams, & Löwe, 2006]) when validated by SADS (Self-reported Anxiety and Depression Symptoms), which is based on the DSM-5 (American Psychiatric Association, 2013) criteria, akin to the structured interview MINI (Sheehan et al, 1998) diagnoses. Freely generated word responses analysed with NLP have been found to correlate with the primary and secondary symptoms in DSM-5 that are associated with Major Depressive Disorder (MDD) and Generalized Anxiety Disorder (GAD; Kjell, Johnsson, Sikström, 2021). QCLA has also been shown to correlate with theoretically relevant cooperative behaviour, especially for prosocial individuals in a give-some dilemma game, whereas rating scales were unable to show the expected effect (Kjell, Daukantaitë & Sikström, 2021). Finally, QCLA allows for description of mental constructs, for example, on the group level by visualizing in word clouds what words are associated with high or low degree of depression (Kjell et al, 2019), or on the individual level where words such as *separated, alone*, and *unemployed* provide further understanding of a potential patient than what is communicated with rating scales.

In summary, these studies suggest that QCLA may have a considerably high accuracy and thus have the potential to rival the measurement validity of rating scales. However, few studies have compared the ecological validity of language responses compared to rating scales. Given the current trend of research finding language to be a promising and accurate measure of mental states, we were interested in how respondents perceive language-based response formats, including free texts, descriptive words, and selected words from a predefined list. We investigated whether respondents with or without diagnoses and clinicians view these language-based response formats to be more *precise* in communicating their mental states and whether rating scales are perceived as easier to respond to.

Given the recent progress in the accuracy of computational methods for analysing text responses, and that language is the natural way for people to communicate their mental states, we hypothesize (H1) that respondents view free text as being more *precise* in communicating their mental states compared to rating scales. In contrast, we hypothesize (H2) that rating scales are *easier* to respond to than free text responses. Furthermore, we hypothesize (H3) that formats related to keywords have ratings in between the free recall and rating scales formats. Finally, because language is the natural way for people to communicate their mental state, we hypothesize (H4) that respondents are more likely to prefer free text responses while communicating with a clinicians as compared to what clinicians believe that the respondents prefer.

We tested these hypotheses by first letting respondents answer questions related to depression. The choice of depression, rather than other mental disorders, was governed by the fact that it is a common mental disorder, and that our research group has successfully focused on measuring depression using NLP in previous studies (e.g., Kjell et al, 2019). We used four response formats – free text, descriptive words, selected words, and rating scales – where the order of the listed formats went from very open to closed. In the analyses we focused primarily on comparing the free text and rating scales, where we expected the descriptive and selected words condition to have intermediate rating scores. The choice of these four formats was based on the fact that we previously have been using them in earlier studies (e.g., Kjell et al., 2019; Kjell, Sikström, Kjell, Schwartz, 2021). We then asked the respondents to rate their view of the response format on the following 12 dimensions. How precise it was to communicate their mental states with the response format was measured by eight questions about *precision* in communication, the *nature* of their mental states, evoking or *reinstating* emotions, how they can *elaborate* their answer, how they can *think* more, how the format provides opportunities to consider the construct in *different* ways, if it is a *natural* way to communicate mental states, and how well they can *relate* to communicating with the format. How easy it is to respond to the response format was measured by three questions about how *easy, fast*, and *demanding* it was to use. Finally, one item measured the *preference* for communicating using the response format. This study was not preregistered.

## Materials and Methods

*Participants* were recruited from the Prolific website (https://www.prolific.co/). The inclusion criteria were US nationality, with English as first language, and 18 years of age or older.

The clinician condition included participants who stated they were employed as nurse, psychologist, or medical doctor. There were 92 participants on Proflic in this group, where 62 of these participants started the study and 40 completed it.

In the respondents condition (*N* = 304), a pre-screening was conducted to find a population where half of the respondents self-reported that they had an ongoing major depression diagnosis (MDD) or generalized anxiety disorder (GAD) and the other half were respondents who did not report these diagnoses. A total of 145 respondents reported MDD, 103 reported GAD, and 82 reported being diagnosed with both MDD and GAD. The mean age was 32.7 years, ranging from 18 to 74 years, with a standard deviation of 11.5 years. There were 75 men and 224 women, and 1 preferred not to say. To participate in the study were given an incentive of 1.88 pounds.

### Material

In the free text format, respondents were asked to write a descriptive text with a minimum of 20 words and a maximum number of 1000 words. In the descriptive words format, respondents were asked to type five descriptive words (Kjell et al., 2019). In the select words format, the instructions were to choose 5 words from a set of 30 words, where the selection of words was taken from the most commonly used words in previous studies using the descriptive words format (Kjell et al., 2019). Finally, in the rating scales format respondents used a traditional Likert-scale (PHQ-9, Kroenke, Spitzer, Williams 2001) consisting of a set of statements to which the respondents were asked to respond by choosing one of five responses from 0 (Not at all) to 4 (Nearly every day).

Attitudes towards the four response formats were assessed using 12 statements (see the Appendix) created by the authors with the purpose of measuring the precision in communicating mental health, the cognitive easy to respond, and preference for using the formats. The statements related to precision were: *precision* in communication, *reinstating* emotions, allowing one to *elaborate, thinking* things through, thinking about it in a *different* way, *natural* to communicate, and *relatable*. Statements related to cognitive easy included: *demanding, easy*, and *fast* to produce.

Finally, there was one statement for *preference* for communication. Here we chose Likert scales as the response format because this is currently the most accepted way to quantify attitudes and because the QCLA has not been validated for attitudes. There were five alternatives – 0 for “Don’t agree at all”, 1 for “Don’t agree to some extent”, 2 “Neither agree nor disagree, 3 “Agree to some extent”, and 4 “Fully agree”. The scale on the *demanding* question was reversed to make it compatible with responses on the *easy* and *fast* questions, and is therefore henceforth referred to as *less demanding*.

### Procedure

The study consisted of a survey (constructed in LimeSurvey) that was shared with potential respondents via Prolific. Participants were first asked for informed consent to participate in the study and instructed that they could leave at any time. This was followed by instructions for how to answer the survey (see the Appendix). Participation was anonymous. The study was divided into two phases, where Phase 1 was related to the assessment of mental health in different response formats, and Phase 2 was answering questions related to the perception of the response format.

In Phase 1, participants in the respondent condition were asked to answer questions regarding whether they had been depressed during the last two weeks. They were assessed four times, each time with a different response format, namely free text, descriptive words, selected words, and rating scales. The order of the response formats was randomized and there was no time between the tests. Participants in the clinician condition were asked to read the questions so that they understood the response format, but not to respond to the questions. The reason for why clinicians should not respond to the questions was to simulate clinicians’ natural working environment as non-respondents.

Phase 2 collected the 12 preference ratings towards the four response formats used in Phase 1. The order of the statements was randomized. The survey took approximately 15 minutes to complete.

The study was conducted in accordance with the Swedish Ethical Review Act (SFS 2003:460). The data is available upon request from the corresponding author.

## Results

### The respondent condition

The results are summarized in Table 1a for the respondents and in Table 1b for the clinicians. Figure 1 shows the respondent condition using the rating scales as the baseline. Free text and rating scale responses were compared with two tailed paired t-tests.

**Table 1.** Means and standard deviations of attitudes for respondents and clinicians

a) Respondent condition
| Questions | Rating Scale | Free Text | Descriptive Words | Selected Words | *p* |
| --- | --- | --- | --- | --- | --- |
| Elaborate | 0.98 (1.14) | 3.27 (1.02) | 1.64 (1.10) | 1.44 (1.20) | <.0001 |
| Precise | 2.11 (1.23) | 3.05 (1.00) | 2.29 (1.12) | 2.23 (1.18) | <.0001 |
| Think | 1.66 (1.28) | 2.45 (1.35) | 1.79 (1.27) | 1.79 (1.27) | <.0001 |
| Reinstate | 1.35 (1.17) | 2.10 (1.34) | 1.59 (1.21) | 1.50 (1.20) | <.0001 |
| Nature | 2.37 (1.10) | 3.03 (1.07) | 2.38 (1.05) | 2.29 (1.09) | <.0001 |
| Different | 1.09 (1.08) | 1.39 (1.20) | 1.35 (1.16) | 1.39 (1.19) | <.0001 |
| Natural | 2.40 (1.19) | 2.53 (1.35) | 2.04 (1.21) | 2.15 (1.24) | .2060 |
| Relate | 2.69 (1.04) | 2.81 (1.13) | 2.45 (1.00) | 2.46 (1.00) | .1460 |
| Prefer | 2.44 (1.00) | 2.52 (1.13) | 2.16 (0.96) | 2.15 (0.97) | .4450 |
| Easy | 3.07 (1.00) | 2.21 (1.28) | 2.15 (1.17) | 2.30 (1.20) | <.0001 |
| Less demanding | 2.92 (1.04) | 1.71 (1.3) | 1.99 (1.17) | 2.12 (1.18) | <.0001 |
| Fast | 3.06 (1.04) | 1.81 (1.24) | 2.02 (1.09) | 2.23 (1.10) | <.0001 |

b) Clinician condition
| Questions | Rating Scale | Free Text | Descriptive word | Selected Words | *p* |
| --- | --- | --- | --- | --- | --- |
| Elaborate | 1.05 (1.17) | 3.21 (0.91) | 2.09 (1.02) | 1.67 (1.25) | <.0001 |
| Precise | 1.93 (1.12) | 2.86 (1.08) | 2.51 (1.08) | 2.56 (1.26) | <.0001 |
| Think | 2.02 (1.14) | 2.77 (1.32) | 2.28 (1.12) | 2.07 (1.10) | 0.0006 |
| Reinstate | 1.70 (1.01) | 2.47 (1.39) | 2.07 (1.12) | 2.30 (1.08) | 0.0044 |
| Nature | 2.53 (0.96) | 2.67 (1.17) | 2.81 (0.85) | 2.67 (0.87) | 0.0018 |
| Different | 1.70 (1.17) | 1.79 (1.26) | 1.60 (1.09) | 1.88 (1.20) | 0.5334 |
| Natural | 2.35 (1.21) | 2.33 (1.29) | 2.58 (0.98) | 2.47 (1.12) | 0.6518 |
| Relate | 2.47 (1.01) | 2.28 (1.10) | 2.63 (0.82) | 2.70 (0.89) | 0.9311 |
| Prefer | 2.84 (0.87) | 1.44 (1.16) | 2.28 (1.01) | 2.47 (1.10) | 0.4302 |
| Easy | 2.95 (1.07) | 1.56 (1.31) | 2.21 (1.08) | 2.47 (1.24) | <.0001 |
| Less demanding | 2.66 (1.08) | 0.79 (0.99) | 1.63 (1.00) | 2.12 (1.07) | <.0001 |
| Fast | 2.77 (1.00) | 1.00 (1.09) | 1.95 (0.87) | 2.26 (1.05) | <.0001 |

*Note.* The table shows mean values with standard deviations in parentheses. The *p*-values show whether rating scales differed significantly from the free text values using two tailed paired *t*-tests. The rows are ordered after their ratings in the free text minus the rating scale condition in the respondent condition.

**Figure 1.**
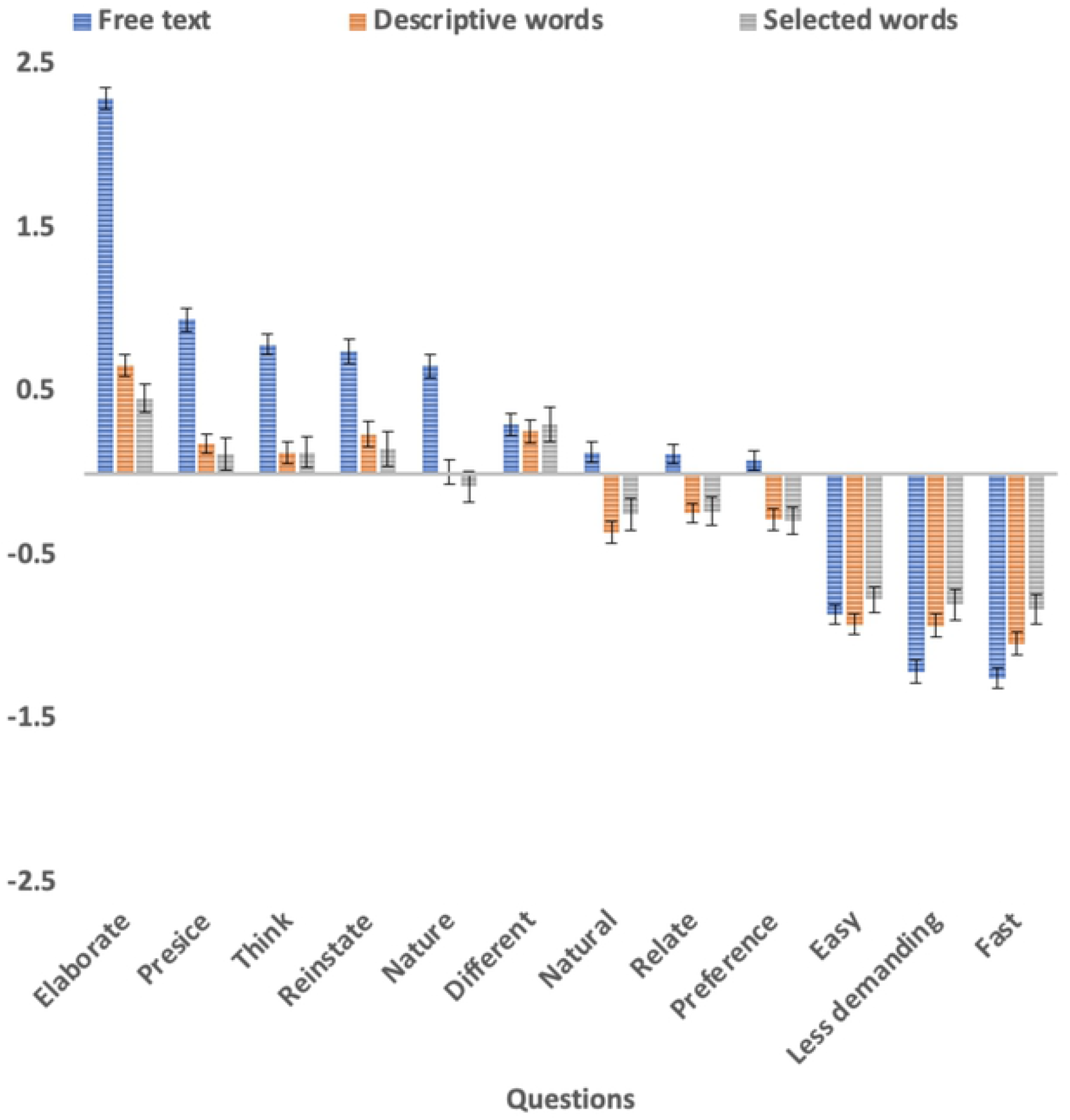
Free text, descriptive words, and selected-words responses using rating scales as the baseline for the respondent condition. *Note*. The y-axis shows the mean response and standard errors for free text (blue), descriptive words (red), and selected words (grey) minus the mean response for rating scales. The x-axis is ordered by the free text minus rating scales value.

The respondents viewed the free text as being more precise than the rating scales (Hypothesis 1) for 6 of the 9 conditions: more *precise* (*t*(302) = 8.16, *p* < .0001, *d*’ = .88), better in *reinstating* (*t*(302) = 6.28, *p* < .0001, *d*’ = .62) their emotions and expressing the *nature* (*t*(302) = 5.31, *p* < .0001, *d*’ = .54) of their mental state, *elaboration* (*t*(302) = 20.87, *p* < .0001, *d*’ = 2.0), and deeper *thinking* (*t*(302) = 6.20, *p* < .0001, *d*’ = .62), and made them think in a *different* way (*t*(302) =2.63, *p* < .0001, *d*’ = .26) compared to the rating scales. However, three measures showed no significant (*p* > .05) difference between free text and rating scales: how *naturally* respondents could *relate* to answering the questions in the response format or to what extent they would *prefer* to respond in a format for a clinician.

The respondents rated the rating scales as easier than the free text response formats (Hypothesis 2) in the three measures aimed to capture this: rating scales were perceived as *easier* (*t*(302) = – 7.62, *p* < .0001, *d*’ = –.67), *faster* (*t*(302) = –11.70, *p* < .0001, *d*’ = –1.13), and less *demanding* (*t*(302) = –9.95, *p* < .0001, *d*’ = 1.03). The objective time to complete the rating scale (*m* = 60.9 s) was also significantly faster than the time to complete the free text response (*m* = 186.3 s, *t*(302) = 10.0, *p* < .0001, *d*’ = .58). The selected words condition (*m* = 50.7, t(302) = –3.41, *p* < .0001, *d*’ = -.20) was somewhat faster than the rating scales, whereas the descriptive words condition was slightly slower (*m* = 77.3, *t*(302) = 3.52, *p* < .0001, *d*’ = .20).

Consistent with Hypothesis 3, the descriptive words and the selected words formats had results that were nominally in between the free text and the rating scales in all conditions with a significant difference between the rating scales and free text responses.

### Clinician condition

The results in the clinician condition were consistent with the results in the respondents’ conditions in all measures, except in ‘thinking about it in a *different* way’ ratings where there was no significant difference between free text and the rating scales condition.

All significant findings in the respondents’ conditions, following correction for multiple comparisons (*N* = 12), had *p*-values < .001, and for non-significant *p* > .05 alpha was set to .05.

### Comparing respondents’ and clinicians’ conditions

Respondents had significantly stronger attitudes (Hypothesis 4, see Figure 2A) for free text responses over rating scales compared to clinicians for *preference* (*t*(345) = 5.25, *p* < .0001, *d’* = 0.85) following correction for multiple comparisons, and for the following four other attitudes without correction for multiple comparisons: *less demanding* (*t*(345) = 2.59, *p* = .0099, *d’* = 0.42), *easy* (*t*(345) = 2.11, *p* = .0353, *d’* = 0.34), communicate the *nature* of their mental state (*t*(345) = 2.22, *p* = .0269, *d’* = 0.36), and *fast* (*t*(345) = 1.97, *p* = .0491, *d’* = 0.32).

**Figure 2A.**
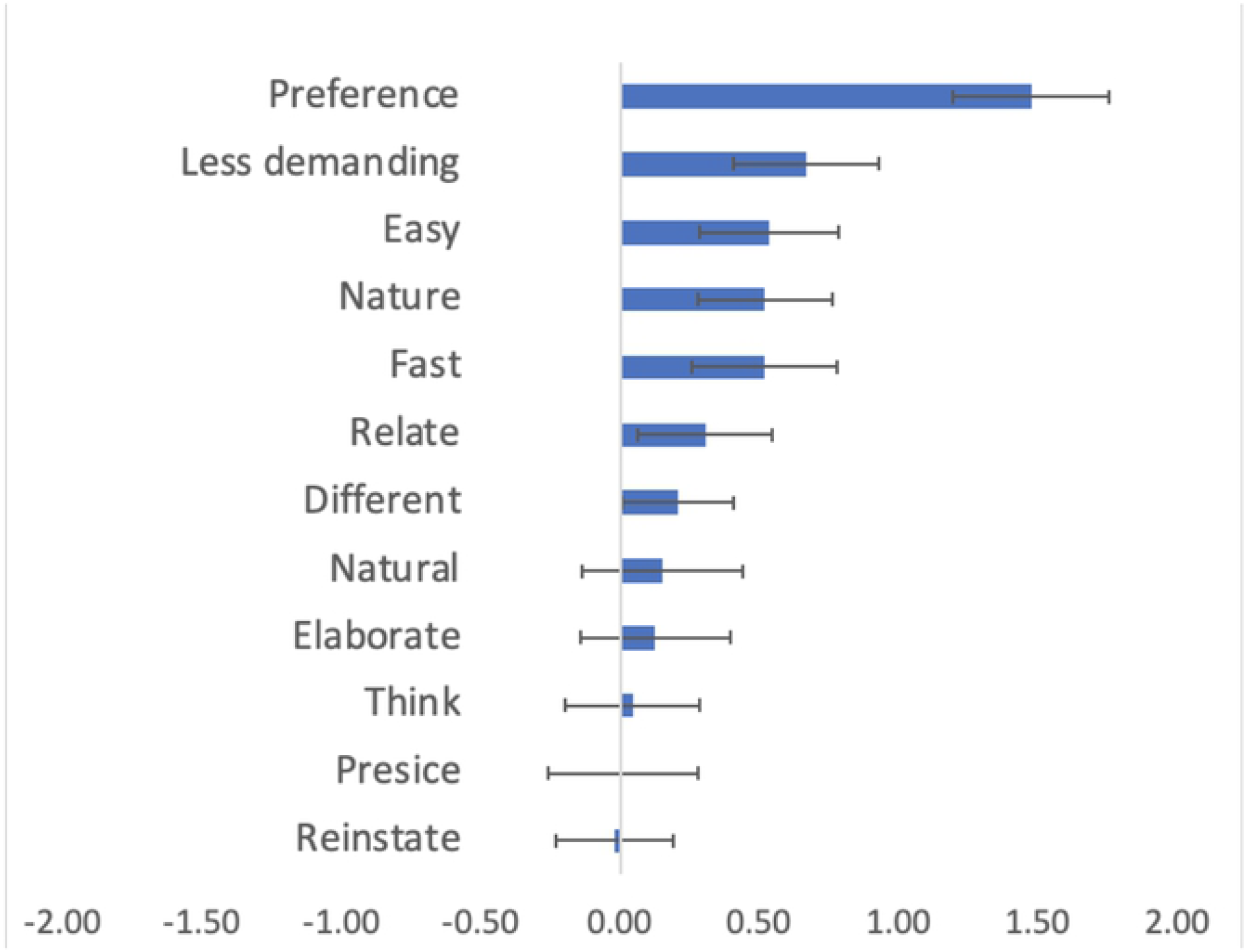
Respondents’ preferences of free text responses over rating scales using clinician as the baseline

**Figure 2B.**
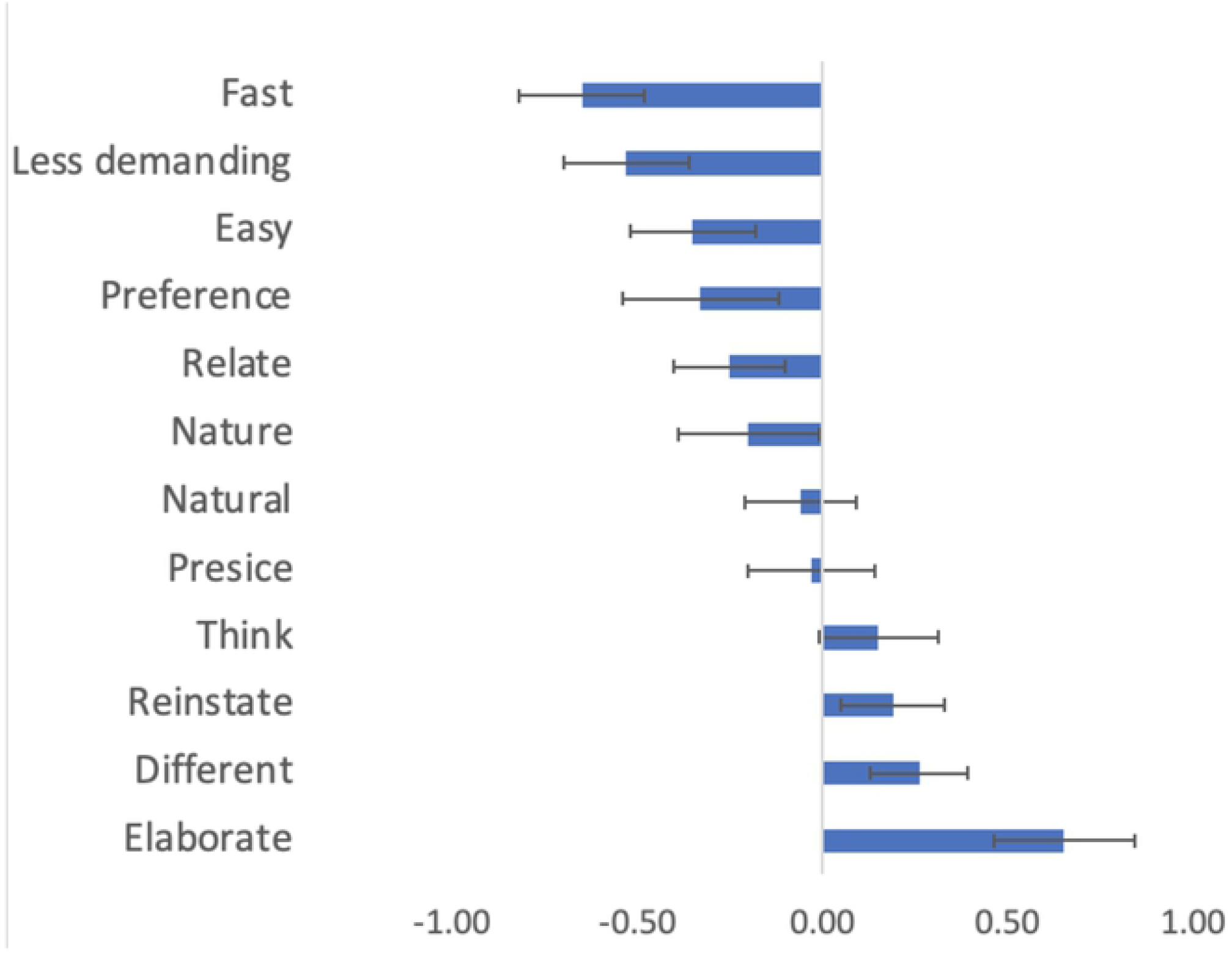
Normal controls’ preferences of free text responses over rating scales using diagnosed as the baseline *Note*. The figure shows the mean ratings of free recall responses minus the mean ratings for rating scales. Figure 2A shows respondents minus clinicians where a positive value indicates that respondents prefer free text responses over rating scales more than clinicians do. Figure 2B shows healthy controls minus participants diagnosed with depression or anxiety, where positive values indicate that healthy control prefer free text response over rating scales more than diagnosed do. The error bars are standard errors.

### Comparing patients with normal controls

The normal control rated a higher advantage in elaboration for free text responses over ratings scales compared to patients with self-reported ongoing MDD or GAD (*t*(329) = 3.51, *p* = .0005, *d*’ = 0.39), whereas patients reported a larger advantage of faster responses rating scales compared to free text responses (*t*(329) = -3.79, *p* = .0002, *d*’ = -0.42) following correction for multiple comparisons (see Figure 2B).

## Discussion

The results clearly show that respondents and clinicians viewed the free text response format as more precise for communicating their state of mind compared to rating scales. This finding is consistent with recent work showing that language-based responses, analysed by state-of-the-art computational language models, show high accuracy and are competitive with and sometimes outperform rating scales (Kjell et al., 2019; Kjell et al., 2021; Kjell et al., 2021, Sikström et al., in progress). Together these findings open up possibilities of using open-ended responses without sacrificing accuracy in measuring different constructs.

Although our data do not allow causal claims, we speculate that one reason for why text responses may be perceived as being more accurate is that they allow more specific *elaborations* of the respondents’ state-of-mind, making them think more about the construct. In addition, our data show that the respondents rated free text responses as enabling them to view the construct in a different way, and previous research suggests that this may be an important aspect for improved mental health (Pennebaker & Chung, 2007). The results did not show any difference between the free text and rating scales in the dimensions of being *natural*, easier to *relate* to, or more *preferred*. A possible reason for this is that *natural* and *relate* are concepts that may be interpreted in different ways, where open formats are less constrained and therefore harder to connect to these constructs.

The results also showed that both respondents and clinicians rated rating scales as *faster, easier*, and *less demanding* than free text responses, and where the rating scales also took less time to perform than free text responses. The fact that rating scales are easier is obviously an advantage; however, this may be contrasted to the finding that they are rated as less precise. A possible interpretation of this is a trade-off between speed and accuracy, where the rated precision of free recall responses can be traded with slower response (Wickelgren, 1977). Furthermore, the selected-words responses were actually answered faster than the rating scale, whereas the descriptive words were answered just slightly slower than the rating scales. Currently, rating scales are the most common response format in Prolific (Flake, Pek, & Hehman, 2017), but the speed of responding to the open-language questions may increase as the respondents become more used to these response formats.

Respondents had the same *preference* for communicating their mental states to a clinician using rating scales, whereas the respondents indicated a stronger preference for free text formats compared to the clinicians ratings. Interestingly, this was also the rating where the difference between respondent and clinician ratings was the largest (*d’* = 0.85). Furthermore, clinicians exaggerated how slow, demanding, and hard (as measured by *fast, less demanding*, and *easy*) the respondents rated free recall responses. Thus, on several questions clinicians could not estimate how respondents rated the questions related to the response formats on depression. The findings cannot easily be explained with the possibility that the clinicians have little knowledge of the respondent’s point of view, as they could also have simulated the respondents answer with their own attitudes. An alternative explanation is that the clinicians’ attitudes are different from the respondents.

Another difference is that the respondents had the opportunity to answer the four response formats related to depression, which the clinicians were instructed not to do. This design was intentionally setup to simulate the natural situation where respondents respond to questions, whereas clinician do not. We argue that this setup is more interesting to study as it is closer to the real-life interaction between respondents and clinician, compared the alternative design where clinicians and the respondents would have been treated identically.

Free text response is a more open format compared to freely generated descriptive words or selected words. We therefore expected that the preferences for the descriptive word responses would show results that were in between the free text and rating scales, which also was found in the majority of the questions.

The findings that participants rate free text response higher than rating scales on several dimensions related to precision, can be related to recent advances in assessment of depression and anxiety using computational methods. For example, Kjell et al. (2019; Kjell, Sikström et al., 2021) used natural language processing algorithms (e.g., Latent Semantic Analysis [LSA]; Landauer, 1997; BERT, Devlin, 2019) to map the text responses to vectors representing participants description of their mental health. They then used Machine Learning (ML, multiple linear regression) to build a model to predict ratings scales of mental health (e.g., PHQ-9 and GAD-7). The results showed that text responses can be accurately used to predict rating scales. Advantages with this method is that it provides an accurate and standardised procedure to assess mental health in the format that is the natural way for participants (patients) to communicate their mental health. Disadvantages with this method includes that the participant and clinicians need to understand and feel trust in the results. These computational findings suggests that text data, consistent with participants ratings, can be used to make quantitative assessments of mental health with high accuracy.

### Limitations

Although, the study showed clear significant results, the number of participants in the clinician condition was rather limited, despite the fact that we invited all available clinicians on Prolific given inclusion/exclusion criteria. The limited size of participants did not allow us to control for various demographic factors such as age, gender, education, reading level, cognitive ability, personality profiles, etc. Furthermore, the data only includes measures related to depression, whereas other mental health issues such as anxiety, post-traumatic stress disorder, stress, etc, are not covered. Therefore, we cannot make any claims whether the findings generalized to other disorders than depression.

### Other remarks

The usage of open-ended text responses require other demands than rating scales. For example, as open-ended questions are more demanding, less people tend to answer them (Zhou et al., 2017). Furthermore, the type of questions matter, where comment-specific questions (e.g., “I feel happy to meet her”) yield higher response rates than explanatory-specific questions (e.g., “I meet her because she makes me feel happy”). Response rates also depend on demographic variables, where young males with high online literacy are more likely to answer open-ended questions than old females with low online experience (Zhou et al., 2017). Finally, the format of free text response box for free text response matters, where more lines evoke longer text responses (Spörrle et al., 2007).

## Conclusion

Our finding suggests that respondents view free text responses as more precise than rating scales, possibly because they allow better opportunities for elaboration. Progress in computational language assessment now provides opportunities to quantify languages with an accuracy that matches respondents’ expectation of high precision and validity while using text response as outcome variables in behavioural science. The results suggest that future assessment of mental health patients may benefit from support by computational methods that uses open-ended text data.

## Data Availability

The data includes texts describing personal information of mental health and can therefore not be shared due to ethical reasons.

## Appendix

### Instructions to respondents

In this study we are interested in what you think about different responses formats when describing your level of depression. First you will be asked to answer four types of questions about depression with different response formats, including: The response formats are: filling out a *Rating* scale, *Selecting* 5 descriptive words from a list of 30 words, writing 5 descriptive *Words*, and writing a *Text* of 20 to 1000 words. Second, you will be asked a series of questions about how you perceived answering with the different response formats. Try to weigh the strength, and the number of aspects, so that it reflects your overall personal level of depression, or lack of depression.

### Instruction to researchers

In this study we are interested in what you think about different responses formats when participants describe their level of depression. First you will view questions about depression with four different response formats. Your task is to look at the questions and response formats so you will understand the questions in the second part of the study. The response formats are: filling out a *Rating* scale, *Selecting* 5 descriptive words from a list of 30 words, writing 5 descriptive *Words*, and writing a *Text* of 20 to 1000 words. Second, you will be asked a series of questions about these response formats.

The questions in Phase 2 for respondents (a) and clinicians (b) are grouped after how they are connected to the hypothesis.

Hypothesis 1: Precision

1 a. How much did the response format allow you to **elaborate**?
b. How much do you think the response format allowed the respondents to elaborate?
2 a. How **precisely** could you communicate your true feelings and symptoms in the following response formats:
B. How precisely do you think the respondents could communicate their true feelings and symptoms in the following response formats:
3 a. To what extent did the response format make you **think** through your problems related to depression?
b. To what extent do you think the response format made the respondent think through their problems related to depression?
4 a. To what extent did the response format evoke or **reinstate** emotions?
b. To what extent do you think the response format evoked or reinstated emotions in the respondents?
5 a. How well could you communicate the **nature** of your depression, or lack of depression, in the following response formats?
b. How well do you think the respondents could communicate the nature of their depression, or lack of depression, in the following response formats?
6 a. To what extent did the response format make you think about your depression in a **different** way?
b. To what extent do you think the response format made the respondent think about their depression in a different way?
7 a. How **natural** was it to communicate your feelings using the following response formats?
b How natural do you think it was for the respondent to communicate their feelings using the following response formats?
8 a. How well could you **relate** to answering the question in the following response formats:
b. How well do you think the respondents could relate to answering the question in the following response formats:

Hypothesis 2: Easy

9 a. How **easy** was it to respond using the following response formats:
b. How easy do you think it was for the respondents to respond using the following response formats:
10 a. How **demanding** was it to respond in the following response formats:
b. How demanding do you think it was for the respondents to respond in the following response formats:
11 a. How **fast** could you respond in the following response formats:
b. How fast do you think the respondents could respond in the following response formats: Hypothesis 3: Preference
12 a. Given you feel very depressed, to what extent would you **prefer** to use the following response format while communicating with a clinician?

